# Deep brain stimulation for post-stroke pain, cognitive and motor function rehabilitation: A comprehensive systematic review and meta-analysis on a novel approach

**DOI:** 10.1101/2025.06.03.25328922

**Authors:** Hung Huu Phan, Aishwarya Koppanatham, Rawan Nawas, Shree Rath, Uneeb Ullah Khan, Imad Akbar

**Author notes:** **Corresponding author:** Phan Huu Hung Department of Medicine University of Medicine and Pharmacy 217 Hong Bang Street, District 5 Ho Chi Minh city, Viet Nam.

## Abstract

**Background:** Deep brain stimulation (DBS) is a neuromodulation technique used to treat various refractory conditions relating to movement disorders. It’s application in the stroke population has shown increasing interest in recent years, introducing promising initial results as a potential therapeutic modality for post-stroke rehabilitation.

**Objectives:** This systematic review and meta-analysis critically examine the efficacy and safety of DBS for post-stroke pain, cognitive and motor outcomes.

**Methods:** The study was registered in PROSPERO (Registration number CRD42024586415). Following PRISMA protocol, we systematically searched PubMed, EMBASE, CINAHL and Cochrane Library databases. The primary outcome was the effectiveness of DBS in improving pain and motor functions after stroke. Secondary outcomes were the effectiveness of DBS in improving cognitive function and post-intervention complications.

**Results:** The final quantitative synthesis included 10 studies reporting 109 patients. The overall pooled 1-year response rate was 0.82 [95% CI: 0.65 - 0.92] (p = 0.03), long-term response rate was 0.71 [95% CI: 0.32 - 0.92] (p < 0.01) on the random effects model. Reported overall procedural-related adverse events was 7% [95% CI: 0.04 - 0.15] and efficacy on chronic post-stroke pain VAS/BPI reduction was from 8.24 [95% CI: 7.55 - 8.72] at baseline to 4.60 [95% CI: 3.63 - 5.57], respectively (p < 0.01). A single-individual study on motor function recovery showed promising initial phase I trial results, with significant functional motor recovery and low rates of adverse events at 9%. Procedural-associated mortality was either not reported or not observed.

**Conclusions:** DBS is a promising and novel approach for post-stroke deficits rehabilitation. With prospective initial results, further high-quality randomized trials are needed to conclusively establish efficacy across diverse post-stroke conditions, particularly for motor function recovery.

**Key points:**

- The study demonstrated a high 1-year response rate of 82% and a long-term response rate of 71%, defined as at least a 30% improvement in symptoms. DBS showed significant efficacy in alleviating chronic post-stroke pain (CPSP), improving motor functions, and enhancing cognitive outcomes.
- Deep Brain Stimulation exhibited a low adverse event rate of 7%, with no procedural-related mortality reported during an average follow-up period of 29.2 months.
- Despite promising results, the study emphasized the need for larger, high-quality randomized controlled trials to validate findings, particularly for motor recovery. Current evidence is still very limited, as reported by early-stage trials. Further exploration of clinical feasibility, cost-effectiveness, and quality-of-life impacts should be highly considered.

## INTRODUCTION

Stroke remains one of the leading causes of mortality and a significant contributor to the global healthcare burden^1,2^. Despite early diagnosis due to its apparent clinical manifestations, delays in time-to-treatment for newly onset stroke continue to pose substantial challenges to effective intervention^3^. Thus, a high prevalence of lifelong post-stroke disability persists, often with minimal improvements in functional capabilities, mental capacity, or quality of life^4^. Efforts in rehabilitative care and palliative treatment have been made to improve patient’s quality of life, yet they constitute the increasing burden for healthcare. Alongside traditional stroke rehabilitation, a multidisciplinary approach has highlighted the importance of incorporating occupational, physical, and interventional therapies to enhance patient outcomes and reduce dependence on lifelong medical care and daily assistance.

Neuromodulation consists of a wide range of interventional techniques, aimed at recovering lost neurological capabilities or compensating for compromised brain areas. These include both invasive and non-invasive approaches such as transcranial direct current stimulation (tDCS), vagus nerve stimulation (VNS), transcranial magnetic stimulation (TMS), spinal cord stimulation (SCS), and deep brain stimulation (DBS)^5^. By affecting various sites of lesions in stroke survivors, these therapeutic approaches offer a synergistic approach alongside traditional rehabilitation. Neuromodulation therapies have been employed to treat patients with post-stroke pain, tremors, hemiparesis, aphasia, cognitive function, and motor deficits^6–14^. Advancements in neuromodulation are shifting the focus of post-stroke care to providing innovative recovery strategies for stroke survivors^15,16^.

Deep brain stimulation is commonly used in various neurological dysfunction and neuropsychiatric conditions. The implication of DBS in Parkinson’s disease, dystonia, and mental disorders including obsessive-compulsive disorder and depression has been noted to provide beneficial outcomes in different patient profiles^17–19^. It is an invasive technique where electrodes are surgically implanted into deep brain structures, such as the thalamus, globus pallidus, or subthalamic nucleus. These regions involve motor control, pain processing, and cognitive regulation^5,20^. By delivering continuous electrical pulses, DBS modulates abnormal neural activity and promotes neuroplasticity. It is particularly effective for post-stroke tremors, chronic pain, and motor deficits. Cerebellar structures have been noted as the potential target for DBS in stroke survivors with motor dysfunction, as it is involved in communicating and coordinating complex signal pathways with the primary motor cortex, via a disynaptic excitatory pathway that passes through the ventral thalamus. A 2009 rat model study on cerebellar ischemia has demonstrated significant motor function recovery under the effect of contralesional lateral cerebellar nucleus electrical stimulation^21^. Recently, a 2023 randomized phase-I clinical trial by *Baker et al.* reported prominent initial results on motor-impaired stroke patients, by targeting the cerebellar dentate nucleus as the site of stimulation^12^. Therefore, DBS has the emerging potential to enhance neuroplasticity, supporting the restoration of motor functions impaired by stroke^22,23^.

Patients with refractory chronic post-stroke pain (CPSP) also benefit from the implementation of DBS^24–26^. This is a debilitating condition affecting up to 10% of stroke survivors, characterized by persistent pain that is often resistant to conventional treatments. The pain can arise from central post-stroke pain due to lesions in sensory pathways or maladaptive plasticity in central pain-processing networks^26^. Thus, DBS has emerged as a potential therapeutic option for managing refractory CPSP, leveraging its ability to modulate neural circuits involved in pain perception and processing. Several studies have reported positive outcomes in stroke survivors with CPSP with significant improvement in pain perception and quality of life while maintaining a low rate of non-lethal adverse events^27–32^. The thalamus, specifically the ventroposterolateral and the ventroposteromedial nucleus - is a key relay station for sensory information, including nociceptive signals. DBS in these nuclei modulates abnormal pain signals by restoring normal thalamic activity and rebalancing sensory transmission^24,29,31^. Other potential DBS target for CPSP includes the periaqueductal and periventricular gray, cingulate cortex, and motor cortex. Although reported outcomes among different stimulation sites produced inconsistent results, DBS represents a promising intervention for refractory CPSP^27,33–35^.

This systematic review and meta-analysis aim to critically evaluate the current evidence surrounding the application of DBS in stroke rehabilitation. Specifically, the review synthesizes data on its efficacy in alleviating chronic pain, promoting motor function recovery, and enhancing cognitive outcomes.

## METHODS

### Study design

We conducted the systematic review and analysis from September 2024 to October 2024. The study’s protocol was pre-registered and published in PROSPERO (Registration number CRD42024586415). We adhered to the Preferred Reporting Items for Systematic Reviews and Meta-Analysis (PRISMA)^36^ guidelines and reported all stages of the review using PRISMA flowchart [Figure 1]. We aim to address the following gap:

1. What are the current applications and evidence levels of Deep Brain Stimulation (DBS) in stroke rehabilitation?
2. What effect do these strategies have on the outcomes of patients diagnosed with stroke and their reliability, feasibility, and prospects based on current observations?

**Figure 1.**
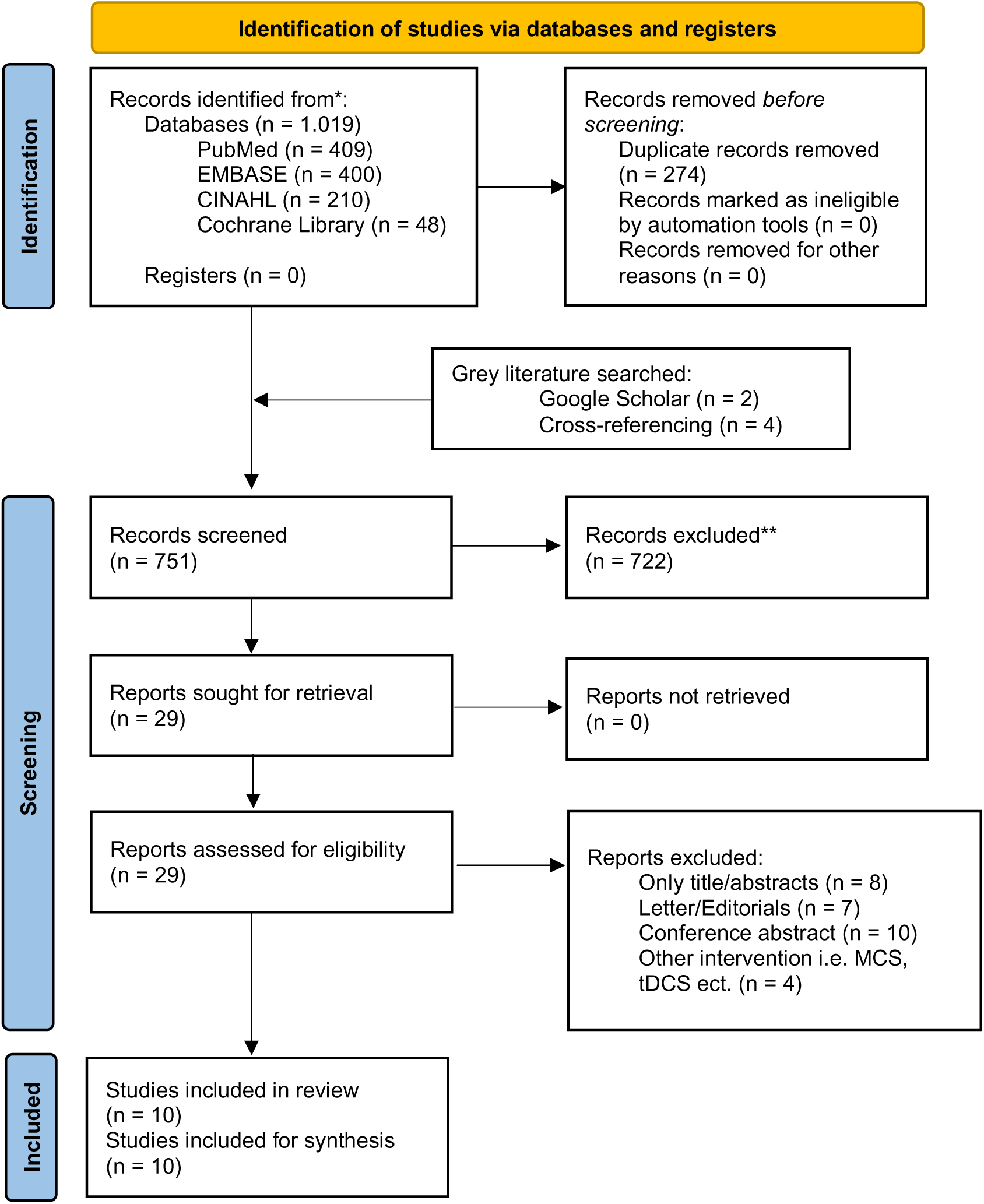
PRISMA flowchart for study selection. **Description:** This flowchart illustrates the PRISMA (Preferred Reporting Items for Systematic Reviews and Meta-Analyses) flow of study selection used in the systematic review of DBS for post-stroke patients. The process details each stage, from the initial identification of records through to the final inclusion of studies.

### Search strategy and information sources

Studies were systematically searched in PubMed, EMBASE, CINAHL, and the Cochrane Library databases up to July 2024, following PRISMA guidelines. To further identify relevant grey literature, article cross-referencing and a Google Scholar search were also conducted. Systematic database searches were performed using a combination of keywords and MeSH terms, with Boolean operators ‘AND’ and ‘OR’ employed to refine the search. Some search terms were truncated with an asterisk (*) to capture multiple word endings. Studies were limited to articles from January 1990 to July 2024 and published in English [Appendix 1].

### Eligibility criteria

Preliminary research yielded a promising application, despite considerable gap in the current literature, regarding the application of DBS in the stroke population. This is consistent with a recent systematic review on the potential of DBS for post-stroke motor recovery, highlighting a need for large randomized clinical trials since current evidence is sparsely reported through individual case reports^22^. For a more comprehensive review and synthesis, we included both observational and interventional studies that align with the eligibility criteria.

We followed the Population, Intervention, Control group, Outcome, Study design, and Time (PICOST) framework for inclusion of suitable studies [Table 1]. To ensure evidence consistency, we excluded abstract-only papers, articles without full-text availability, systematic and scoping reviews, editorials, case reports, animal studies, and studies not reporting on the efficacy of DBS for stroke patients. Articles which provided unreliable, duplicated or inadequate data, or not published in English were also excluded.

**Table 1.** PICOST criteria for study inclusion. **Description:** This table outlines the PICOST (Population, Interventions, Control, Outcomes, Study Design, Time) criteria used to determine the eligibility of studies for inclusion in the systematic review of Deep Brain Stimulation for post-stroke patients.

| Criteria | Determinants |
| --- | --- |
| Population | Adult patients (18 years and older) who have suffered a stroke and present with cognitive and/or motor function deficits. |
| Interventions | Deep Brain Stimulation (DBS) targeting specific brain regions (e.g., subthalamic nucleus, thalamus, or motor cortex) aimed at improving cognitive or motor rehabilitation after stroke. |
| Control group | Standard rehabilitation therapy (e.g., physical therapy, cognitive therapy, or pharmacological treatments) without DBS, or placebo stimulation. |
| Outcome | Primary Outcomes:<br>Improvement in motor function (e.g., using the Fugl-Meyer Assessment scale, Modified Rankin Scale)<br>Improvement in pain scales (e.g., VAS/BPI) and other related stroke sequela<br>Secondary Outcomes:<br>Quality of life (e.g., measured by Stroke Impact Scale or EuroQol-5D)<br>Neuroplasticity markers (if available)<br>Safety outcomes (e.g., adverse events, device-related complications) |
| Study design | Prospective studies including randomized and non-randomized clinical trials, cohort studies, Case control studies and case series ≥ 5 patients |
| Time | Studies published from January 1990 up until September 2024 |

### Study selection

All identified studies were imported into EndNote (version X9), where duplicates were identified and removed. The remaining studies were then exported to Rayyan.ai for blinded title and abstract screening by two independent reviewers (R.N, S.R), who applied predefined inclusion criteria. After the screening phase, the blinded status was lifted, and any disagreements were resolved through discussion with a third reviewer (U.U.K) and further review based on the inclusion criteria. Abstracts that met the initial screening criteria were requested for full-text retrieval and prepared for subsequent review and synthesis. The process was documented and visualized using a PRISMA flowchart [Figure 1].

### Data extraction

Data Extraction was carried out independently by two reviewers using a standardized form to ensure consistency. Extracted data encompassed key study characteristics, including the first author, publication year, country, study design (such as randomized controlled trials and cohort studies), sample size, and follow-up duration. Patient demographics were noted, including age, gender, stroke type, stroke location, and time since stroke onset, along with baseline clinical characteristics. Detailed information on the intervention was also extracted, including the type of Deep Brain Stimulation (DBS) device used, targeted brain regions (e.g., thalamus, subthalamic nucleus, motor cortex), stimulation parameters (such as frequency, pulse width, and voltage), and the duration of the intervention. For studies with control groups, we gathered details on the comparator interventions, such as standard rehabilitation therapies (e.g., physical and cognitive therapy), sham stimulation, or absence of intervention.

Primary outcome was the improvement in motor function, as measured by standardized assessment tools such as the Fugl-Meyer Assessment and the Modified Rankin Scale^37,38^. Pain relief evaluated using pain assessment tools such as the Visual Analog Scale (VAS) and the Brief Pain Inventory (BPI) was also extracted^39,40^. We defined responders as having ≥ 30% of symptom improvement, reflected by subsequent scoring systems. Secondary outcomes included cognitive function improvement, assessed using cognitive scales and quality of life enhancement, using patient-reported outcomes to assess the broader impacts of DBS on daily living and overall well-being. Safety and adverse events, including rates of complications such as infection, device malfunction, and other procedural risks were also assessed.

### Study risk of bias and synthesis

Two independent reviewers evaluated the included studies for various types of bias utilizing the ROBINS-I tool for non-randomized interventional studies and the RoB 2.0 risk of bias tool for randomized studies. We assessed bias due to confounding, selection of participants into the study, classification of interventions, bias due to deviations from intended interventions, missing data, measurement of outcomes, and bias in selection of the reported result of each study accordingly. Each domain was categorized as having a “Low,” “Moderate” or “Serious” risk of bias. Overall evaluation was reported and visualized through the Robvis tool.

### Statistical analysis

Data analysis was performed using R, with random-effects models utilizing the inverse variance method. For clinical outcomes, standardized mean differences (SMDs) along with 95% confidence intervals (CIs) were computed. The SMD quantifies the effect size of the intervention in each study relative to the observed variability, enabling comparisons of pre- and post-intervention data, regardless of the measurement units. The results were presented with 95% CIs, p-values, and corresponding forest plots. Meta-analysis was conducted for outcomes with sufficient available data.

### Assessment of heterogeneity

To visualize the consistency of evidence reported by various studies, we evaluated statistical heterogeneity across studies using the I² statistic, which indicates the proportion of variability in treatment effects due to heterogeneity rather than random chance. The interpretation of I² is as follows: 0-40% suggests low heterogeneity that may not be of concern; 30-60% indicates moderate level of heterogeneity; 50-90% suggests substantial heterogeneity; and 75-100% reflects considerable level of heterogeneity. In addition, a P-value of 0.10 for the Cochran Q test signifies the presence of significant heterogeneity.

## RESULTS

### Search results and study characteristics

The systematic search yielded a total of 1.019 records across the databases, of which 274 were duplicates and subsequently removed. After screening 751 titles and abstracts, 29 full-text articles were further assessed for eligibility. Ultimately, 10 studies including 109 patients in total, met the inclusion criteria and were analyzed in this review^12,19,27–32,41,42^. These studies included 2 randomized controlled trials, and 8 cohort studies [Table 2]. A PRISMA flow diagram is presented for a visual representation of the study selection process. [Figure 1]

**Table 2.** Summary of included studies. (***Abbreviations:*** *RCT – Randomized clinical trail; MCS - Motor Cortex Stimulation; DBS - Deep Brain Stimulation; AIMS - Abnormal Involuntary Movement Scale; TBI - Traumatic Brain Injury; VS/ALIC - Ventral Striatum/Anterior Limb of the Internal Capsule; CPSP - Central Post-Stroke Pain; VPLa - Ventral Posterolateral Thalamus; CPS - Chronic Pain Syndrome; DN - Dentate Nucleus*)

### Risk of bias assessment

Risk of bias across studies was assessed using the RoB 2.0 for randomized ROBINS-I tool for non-randomized studies. Of the studies, 2 RCTs were classified as having a low risk of bias, 3 cohorts as moderate, and 5 cohorts as high risk due to factors such as confounding variables and measurement of outcomes. Detailed bias domains are illustrated in [Figure 2].

**Figure 2.**
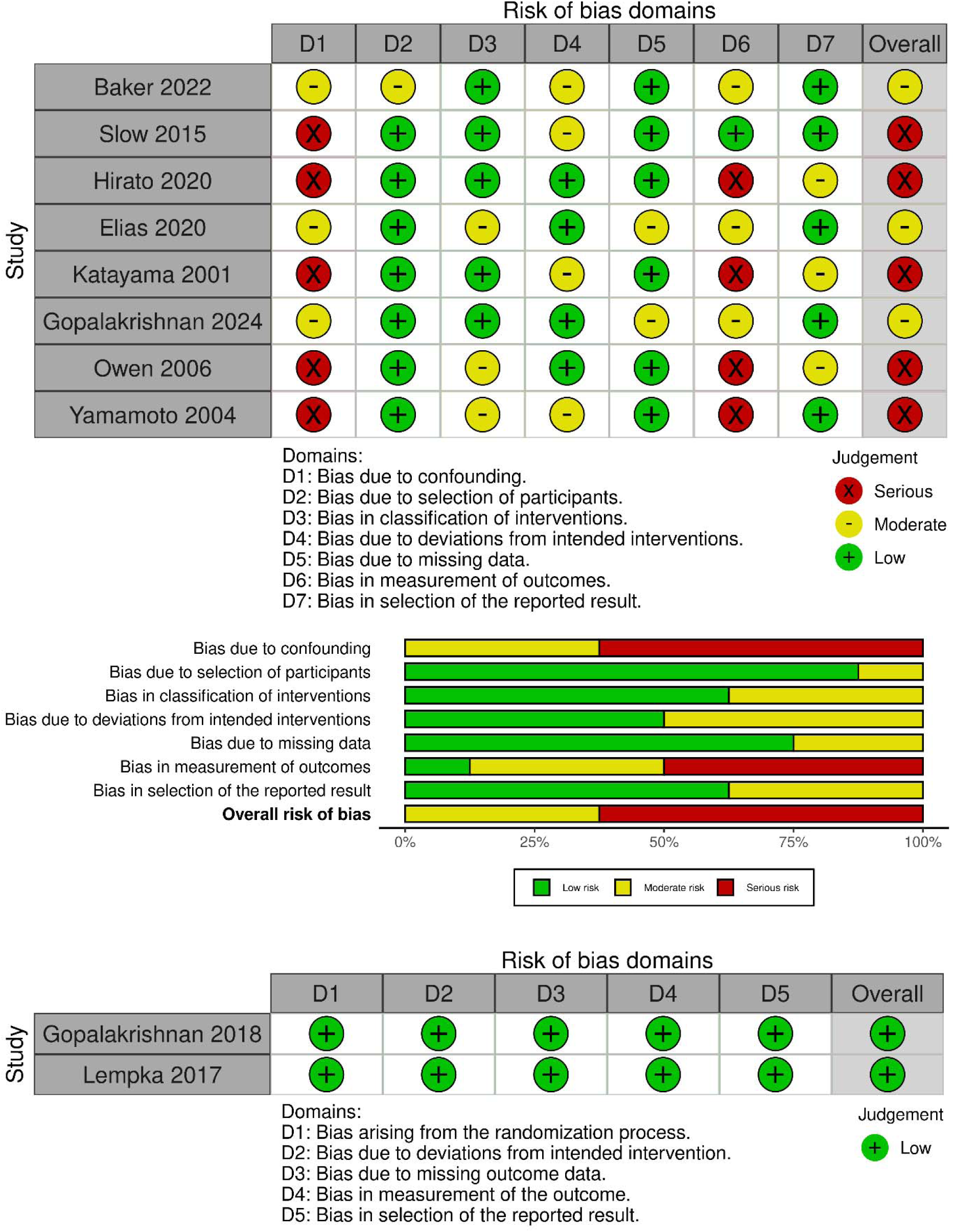
ROBINS-I Risk of Bias assessment for individual non-randomized studies and RoB 2.0 for Randomized studies.

### Population demographics

The included studies covered a broad range of patient demographics and intervention specifics. Across these studies, a total of 109 patients who had experienced post-stroke deficits in pain, cognitive function, or motor function received DBS therapy targeting areas such as the Thalamus (n = 30; 27.5%), Basal ganglia (n = 12; 11.0%), Cortex and Subcortical White Matter (n = 14; 12.8%), Brainstem (n = 11; 10.0%), Cerebellar regions (n = 26; 23.9%), and others. On average, DBS was initiated 6.31 ± 7.78 years after the stroke event, with a follow-up duration of 29.23 [95% CI: 22.45 – 36.01] months, ranging from 6 months^30^ to 6.58 years^29^ [Figure 3C]. The majority of patients had undergone conventional rehabilitation therapies prior to DBS intervention. Mean age was 54.68 ± 11.22 years for the included participants, with 65 (59.6%) male and 44 (40.4%)female.

**Figure 3.**
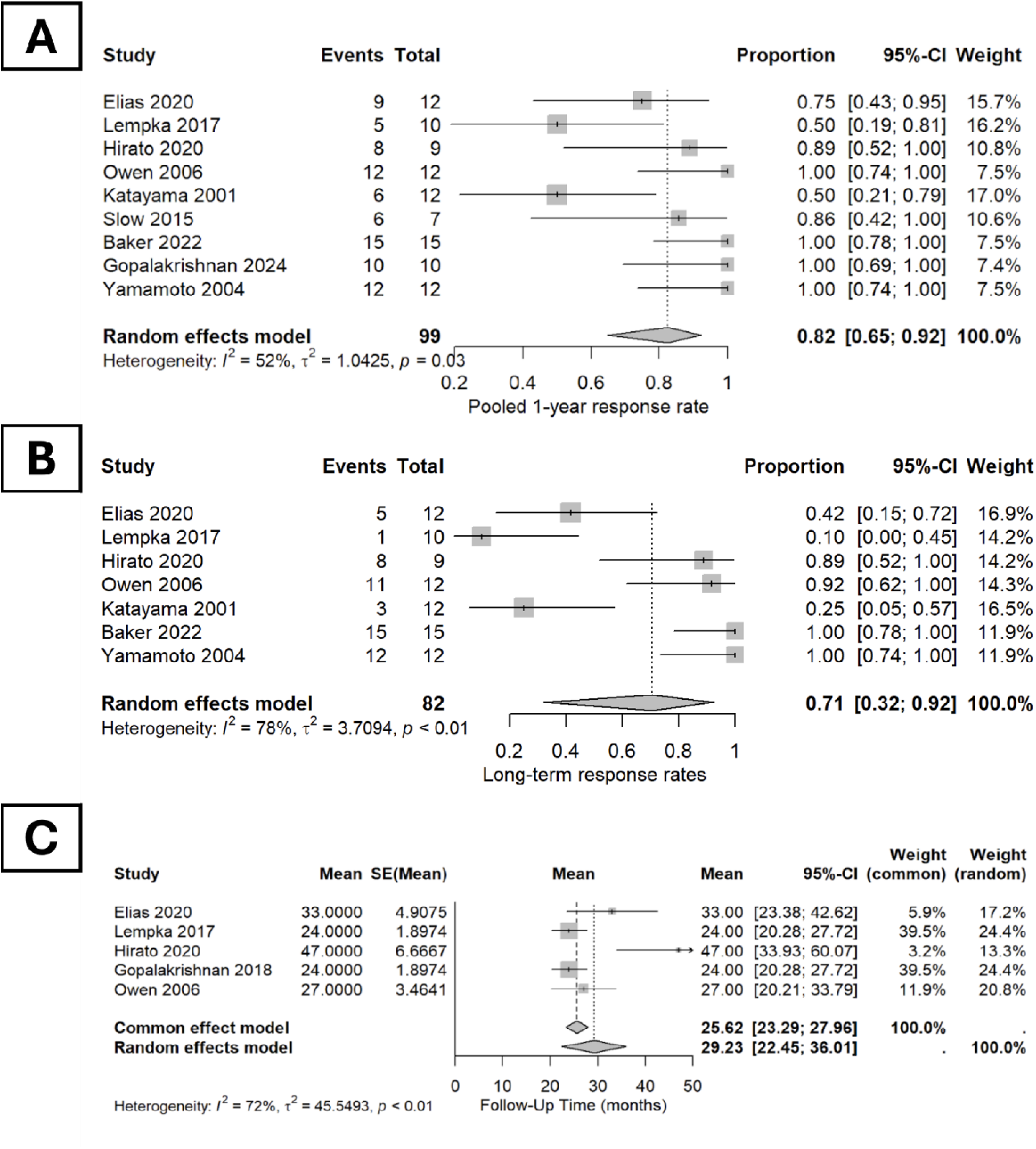
Pooled overall characteristics of included studies. **Description:** This is the forest plot for (A) pooled 1 year response rate, (B) long term-response rate, and (C) follow-up duration - displayed through both common and random effects model. A p<0.05 shows the resutls to be statistically significant.

### Reported outcomes

The overall pooled 1-year response rate on the random-effects model was 0.82 [95% CI: 0.65 - 0.92] (p = 0.03) with moderate heterogeneity (I2 = 52%) and the long-term response rate was 0.71 [95% CI: 0.32 - 0.92] (p < 0.01) with significant heterogeneity (I2 = 78%) [Figure 3A and 3B]. Efficacy on chronic post-stroke pain VAS/BPI reduction was from 8.24 [95% CI: 7.55 - 8.72] at baseline to 4.60 [95% CI: 3.63 - 5.57], respectively (p < 0.01), showing a significant improvement in post-stroke pain [Figure 4A and 4B]. Motor function recovery was observed in several studies^12,41^, with improvements noted on standardized scales such as the Fugl-Meyer Assessment (FMA) and Modified Ashworth Scale (MAS), indicating enhanced motor control and reduced spasticity. Additionally, DBS demonstrated beneficial effects on dystonia and tremors, although these outcomes varied depending on patient characteristics and stimulation parameters^19,42^. However, due to insufficient data, a comprehensive meta-analysis of motor, dystonic, and tremor-related outcomes could not be performed.

**Figure 4.**
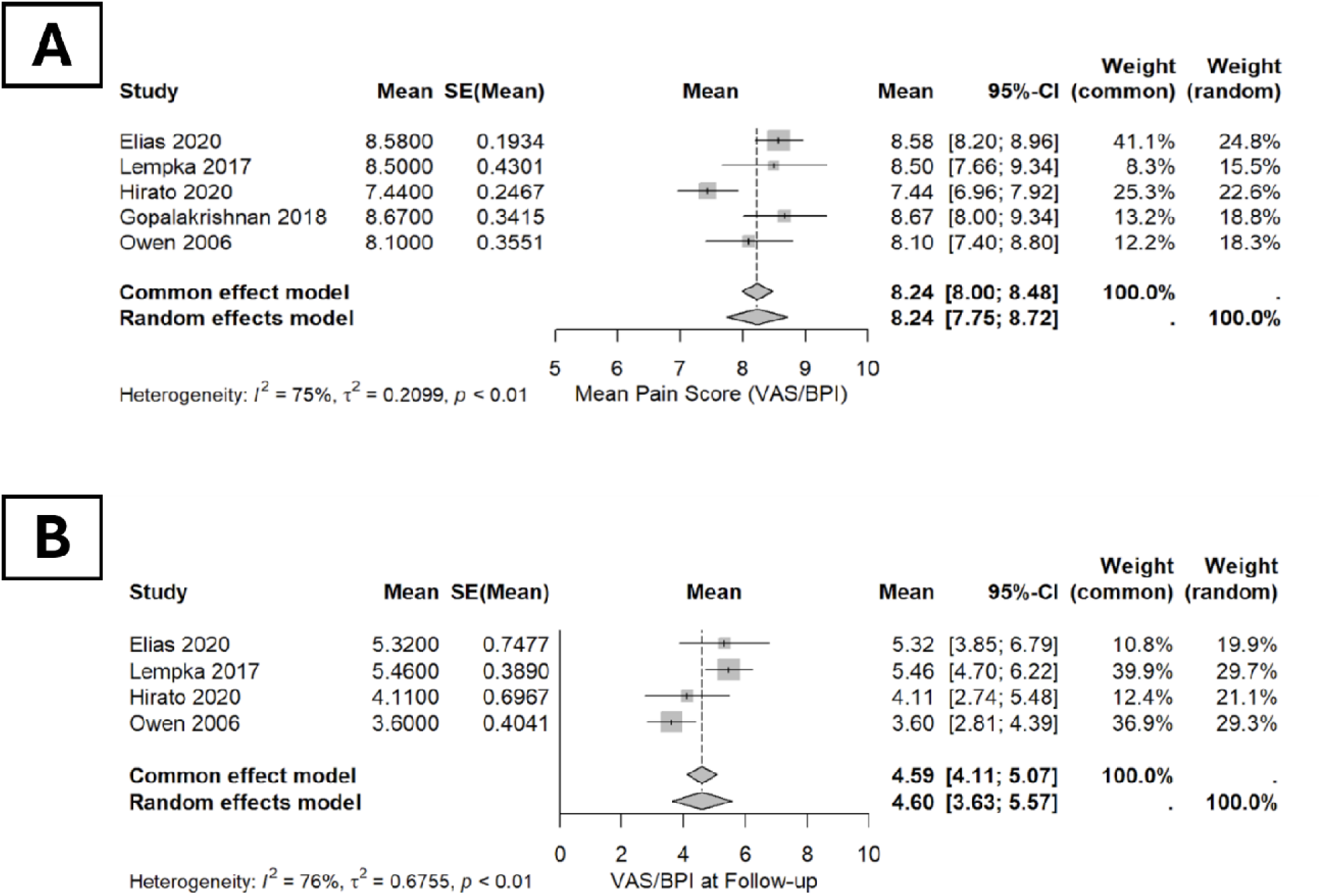
VAS/BPI score at Baseline and at Follow-up. **Description:** This is the forest plot for (A) VAS/BPI pain index at baseline and (B) VAS/BPI pain index at follow-up post-intervention - displayed through both common and random effects model. A p<0.05 shows the resutls to be statistically significant.

Subgroup distribution revealed distinct patterns of brain region involvement across various conditions and underlying etiologies. CPSP accounted for 17 (40.5%) ischemic and 25 (59.5%) hemorrhagic cases. Motor disorders were predominantly ischemic in origin, with 25 cases, while tremors comprising 1 (8.3%) ischemic and 11 (91.7%) hemorrhagic cases. Anatomically, the thalamus was the most frequently targeted region for CPSP (n = 22; 42.3%) and tremor (n = 8; 26.6%), whereas interventions for dystonia were primarily directed at the basal ganglia (n = 5; 66.6%). Notably, motor disorder interventions focused exclusively on the cerebellar region, with 25 cases [Figure 5].

**Figure 5.**
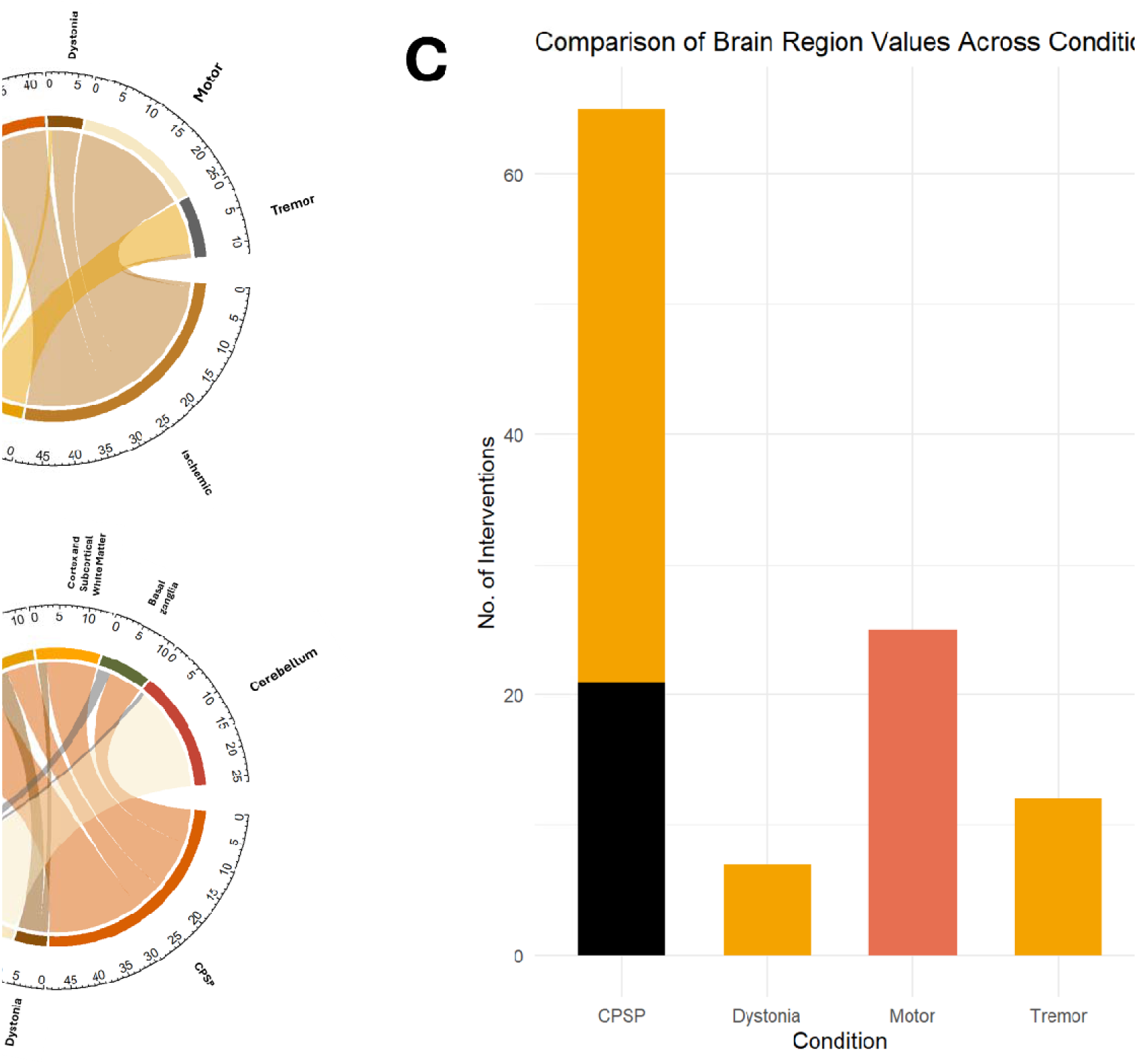
Distribution of Deep Brain Stimulation Target Regions by Clinical Condition in Post-Stroke Rehabilitation. [A] Clinical Condition-Specific Distribution of DBS Targets [B] Anatomical lesions corresponding to Post-Stroke Conditions [C] Anatomical Targeting of DBS for Post-Stroke Conditions. ***Abbreviations:*** *CPSP – Central Post-Stroke Pain; VPL/Vc - Ventral posterolateral/ventral posteromedial nuclei; VS/ALIC - ventral striatum/anterior limb of the internal capsule* **Description:** The image presents three panels (A, B, and C) that illustrate the relationships between neurological conditions, brain regions, and the distribution of interventions. Panel A illustrates the clinical condition-specific distribution of DBS targets using a chord diagram. The diagram connects various conditions such as Tremor, Motor, Dystonia, and CPSP (Central Post-Stroke Pain) to corresponding stroke nature, i.e hemorrahgic or ischemic. Panel B maps the anatomical lesions corresponding to post-stroke conditions. The diagram connects affected regions, including the Cerebellum, Thalamus, Brainstem, and Cortical-Subcortical White Matter, to clinical conditions, highlighting structural damage associated with post-stroke symptoms. Panel C uses a bar graph to represent the anatomical targeting of DBS for post-stroke conditions, comparing the number of interventions across regions. For instance, CPSP is frequently targeted in the VPL/Vc, while Motor symptoms are addressed in the Cerebellar Dentate Nucleus, and Tremor in the VS/ALIC.

Region-specific interventions further highlight condition-specific targeting strategies. The ventral posterolateral/ventral posteromedial nuclei (VPL/Vc) were predominantly targeted in CPSP (n = 44; 69.8%) and tremor (n = 12; 21.4%). In contrast, dystonia interventions were focused on the ventral striatum/anterior limb of the internal capsule (VS/ALIC), with 7 cases. Motor disorders were uniquely associated with the cerebellar dentate nucleus, with 25 interventions. These findings may suggest the importance of targeted interventions to the specific condition and affected brain region, reflecting the nuanced approach required in clinical practice.

### Safety and Adverse Events

Across all studies, DBS was generally well-tolerated, though procedural risks and adverse events were reported. The overall procedural-related adverse event rate was 7% [95% CI: 0.04 - 0.15]., with specific complications including sleep and alertness alterations, headache, post-operative infection, or minor behavioral changes, reported by *Lempka et al. 2017*^28^ and *Baker et al. 2023*^12^. Procedural-associated mortality was either not observed or not reported in the studies, with one noteworthy case of DBS-related seizure which resolved after frequency calibration^28^. This safety profile might suggest the feasibility of DBS as a viable therapeutic option in post-stroke rehabilitation, with manageable risks for most patients.

## DISCUSSION

To the best of our knowledge, this is the single largest study to synthesize long-term outcomes of DBS for stroke survivors. Despite the earliest cohort data being reported since 2001, discussions and practices on this topic remained scarce – only to have gained attention until recently due to its novel application on motor function recovery. There are three key findings presented by this investigation: We found a high long-term response rate of 71%, preceded by an 82% 1-year response rate (defined by at least ≥ 30% symptom improvement). Although we noted a 7% occurrence rate of adverse events, there was no procedural-related mortality over the course of 29.2 months of follow-up on average. There were no suggested differences between demographic characteristics (age at intervention, sex, or disease duration) of responders and non-responders, nor were there sufficient individual patient data (IPD) to infer the effect of technical lead settings on the extent of symptom resolution.

### Clinical feasibility of DBS

In light of whether or not a stroke survivor is a suitable candidate for DBS, the question remains a complex predicament. Neurological recovery potential, following an acute onset of stroke, throughout the chronic phase of rehabilitation, is influenced by many variables^43,44^. The nature of the lesion (i.e. hemorrhagic/ischemic, localization, the integrity of corticospinal and extra-pyramidal tracts, the extent of severity, etc.), accompanied by demographical factors such as age, sex, genetics, and socio-economical status – heavily constitutes a patients outcome^45^. In addition, neuroplasticity is a time-sensitive factor. This suggested that the delay in neuromodulation through rehabilitative efforts might reduce the overall response to DBS, although conflicting findings still exist in the current literature^46–48^.

Stroke-centered DBS requires extensive considerations and strategic clinical assessment. It requires prospective patients to be medically stable, clinically fit, and mentally prepared for DBS, while its nature being a moderately invasive procedure^49^. Potential complications of invasive neuromodulation procedures have been well-reported in the literature: peri-electrode post-surgery edema with subsequent increased intracranial pressure; ill-managed discharge care which can lead to recurrent infections and device malfunctions^50–53^. Cost-effectiveness, patient compliance, and perceived quality of life post-intervention are also a potential topic for investigation. However, there were insufficient data regarding non-procedural related factors that might assist clinical decision-making in practical scenarios.

In patients with conditions refractory to medications and traditional physical rehabilitation, DBS appeared to provide more heightened improvement levels compared to other noninvasive neuromodulation techniques. Meta-analyses of noninvasive stimulation suggested that the improvements achieved through these approaches are generally modest, with a magnitude of 10-30%, and tend to be transient in nature^49,54^. Deep brain stimulation was hypothesized to be more capable of influencing deeper neural structures, being able to stimulate the deep nuclei or white matter tracts. Whereas in motor cortex stimulation (MCS), the confinements of cortical neuroanatomy may inhibit targeted neuromodulatory localization, thus resulting in a transient effect and limited promotion of neuroplasticity^55–57^. In summary, DBS may be particularly useful for moderate to severe neurological deficits localized in subcortical regions, which is also the case for nearly half of pathological presentations in stroke patients^58^.

### Limitations

This study has several limitations that must be acknowledged. The small sample size of 109 patients across 10 studies limits the generalizability and statistical power of the findings. Additionally, substantial heterogeneity in study designs, patient demographics, and outcome measures introduces variability that complicates direct comparisons and synthesis. Only two randomized controlled trials were included, while five cohort studies had a high risk of bias, further reducing the strength of evidence.

Key aspects such as cost-effectiveness and quality-of-life outcomes were not assessed due to the lack of post-discharge characteristics in included studies, limiting the practical applicability of DBS in real-world settings. Moreover, the absence of adequate IPD prevented subgroup analyses to explore factors like stroke type or lesion location. Finally, the considerable heterogeneity observed in some outcomes, such as long-term response rates, highlights variability in reported results. Future multi-center trials with standardized protocols are needed to address these gaps.

## CONCLUSION

Our findings proposed a prospective application of DBS in post-stroke deficit recovery, supported by high long-term response rates, followed by a low-risk procedural profile. Existing clinical data suggested DBS as a valuable approach to managing post-stroke maladaptation, such as persistent central neuropathic pain, tremors, and dyskinesias. Notably, current evidence regarding the exact clinical feasibility of DBS on motor recovery in stroke survivors is still very limited, as reported by early-stage trials. It is therefore important to facilitate more extensive randomized, blinded, and sham-controlled trials to clarify whether longitudinal applications of DBS actually benefit patients, rather than simply producing psychological or contextual influences that may contribute to short-term changes in patient outcomes.

## Data Availability

All data produced in the present work are contained in the manuscript

## ACKNOWLEDGEMENTS

The authors would like to thank the collaborators and colleagues for their contributions throughout the study and express their sincere gratitude to our faculty supervisors for their invaluable guidance and support throughout this research.

## AUTHOR CONTRIBUTION

The first author (PHH): Conceptualized the study, conducted literature review, primary analysis, and drafted the manuscript. The co-author(s): Assisted in data collection and analysis, contributed to writing and editing.

***1. P.H.H:*** Conceptualization, Data Curation, Investigation, Methodology, Project Administration, Writing – Original Draft Preparation, and Writing – Review & Editing
***2. A.K:*** Writing – Review & Editing, Investigation, Data Curation, and Writing – Original Draft Preparation
***3. R.N:*** Writing – Review & Editing, Investigation, and Data Curation
***4. S.R:*** Data Curation, Resources, and Validation
***5. U.U.K:*** Visualization, Writing – Review & Editing
***6. I.A:*** Visualization, Writing – Review & Editing

---

**Appendix 1.** Search term for each database. Search date: 21.09.2024 **Description:** This appendix outlines the search terms used in each database for identifying studies related to the application of DBS on post-stroke patients. The terms were tailored to each database’s search syntax to ensure a comprehensive retrieval of relevant articles.

|  |  |
| --- | --- |
| PubMed | “Deep brain stimulation” AND “Stroke” |
| EMBASE | “Deep brain stimulation” [tiab] AND “Stroke” [tiab |
| CINAHL | “Deep brain stimulation” AND “Stroke” |
| Cochrane Library | "deep brain stimulation" in Title Abstract Keyword AND stroke in Title Abstract Keyword |
| Google Scholar | Deep brain stimulation for post-stroke rehabilitation |

