## Supplementary table 1 for "Deep brain stimulation for post-stroke pain, cognitive and motor function rehabilitation: A comprehensive systematic review and meta-analysis on a novel approach"

| Authors (year) | Design | Sample size | Comparator | Primary outcome | Conclusion |
| --- | --- | --- | --- | --- | --- |
| *Katayama*  *(2001)* | Single arm clinical trial | 45 | Spinal cord stimulation and MCS | Pain control and reduction | Higher pain control achieved in DBS patients as compared to spinal cord stimulation, and lesser than motor cortex stimulation |
| *Yamamoto*  *(2004)* | Single arm cohort study | 15 | N/A | Tremor suppression | DBS led to suppression of tremor within two years |
| *Owen*  *(2006)* | Single arm clincal trial | 15 | N/A | Post-stroke neuropathic pain suppression | DBS improved patient symptoms in 70% of poststroke patients |
| *Slow (2015)* | Single arm clincal trial | 8 | N/A | Resolution of dystonia secondary to a stroke or TBI | The AIMS demonstrated a significant decrease in dystonia severity with an average improvement of 33%. |
| *Lempka*  *(2017)* | Prospective, double-blinded, randomized, placebo-controlled, crossover RCT | 9 | Placebo | Resolution of pain in patients with CPSP | VS/ALIC DBS was safe and effective in addressing the affective component of pain in patients with CPSP. The data suggest that neuromodulation trials may be most effective in reducing suffering by directly modulating the affective components of pain |
| *Gopalakrishnan*  *(2018)* | Double-blinded, crossover RCT | 9 | N/A | Eleviation of chronic pain in patients with CPSP | No significant differences were observed in either measure across the three states. |
| *Elias*  *(2020)* | Retrospective cohort | 17 | MCS | Resolution of CPSP | 12/17 patients (9 DBS) reported benefit during postoperative trial stimulation and proceeded to have their hardware internalized. Only 7/12 patients (5 DBS) were still using and benefiting from their neurostimulator systems |
| *Hirato*  *(2020)* | Double arm cohort study | 9 | N/A | Resolution of CPSP | Adequate and stable pain relief with thalamic VPLa stimulation is obtainable in patients with CPSP who exhibit hyperactivity and electrical instability along the trajectory to this nucleus. Both responders and nonresponders were found to have severe dysfunction of the lemniscal system. |
| *Baker*  *(2023)* | Non-RCT, single arm, phase 1 | 15 | N/A | Safety and feasibility of DBS for CPS motor rehabilitation | Functional gains in patients with lack of adverse effects |
| *Gopalakrishnan*  *(2024)* | Non-RCT, open-label, single arm, phase 1 | 10 | N/A | Motor rehabilitation after a stroke event | DN-DBS combined with rehabilitation was associated with benefits in motor skill and control, with reduced demand for post stroke DN involvement . These benefits were correlated with significant changes in the cortico-cerebellocortical physiology. |

**Table 2. Summary of included studies** (***Abbreviations:*** *RCT – Randomized clinical trail; MCS - Motor Cortex Stimulation; DBS - Deep Brain Stimulation; AIMS - Abnormal Involuntary Movement Scale; TBI - Traumatic Brain Injury; VS/ALIC - Ventral Striatum/Anterior Limb of the Internal Capsule; CPSP - Central Post-Stroke Pain; VPLa - Ventral Posterolateral Thalamus; CPS - Chronic Pain Syndrome; DN - Dentate Nucleus*)
